# Screening for Intracranial Hypertension Using Lighting-Invariant Computational Pupillometry: A Single-Center Pilot Study

**DOI:** 10.64898/2025.12.29.25343108

**Authors:** Kamil Chwojnicki, Magdalena Wujtewicz, Alicja Filipczyk, Rozalia Marszałek-Ratnicka, Julia Czernik, Michal Swiatek, Grzegorz Gramza, Jakub Kukliński, Patrycja Kapica, Hugo Chróst, Marek Dziubiński, Michal Wlodarski, Radosław Chrapkiewicz, Karol Steckiewicz

## Abstract

**Background:** Invasive intracranial pressure (ICP) monitoring is the reference standard for detecting intracranial hypertension but requires neurosurgical expertise and carries procedural risks. Non-invasive methods with high negative predictive value (NPV) could serve as screening tools to rule out dangerous ICP elevations and guide decisions about invasive monitoring. We conducted a pilot evaluation to assess whether a smartphone-based, lighting-invariant quantitative pupillometry platform achieves the high NPV needed for clinical screening.

**Methods:** This prospective single-center pilot study in neuro-ICU enrolled adults with acute neurological emergencies who underwent serial bilateral pupillometry alongside clinically indicated intraparenchymal ICP monitoring. The primary outcome was diagnostic performance of the Pupil Reactivity (PuRe) Score for identifying intracranial hypertension (ICP ≥ 20 mmHg), with emphasis on NPV. Secondary analyses examined PuRe Score correlations with ICP, clinical phenotypes across reactivity strata, ambient lighting invariance, and subgroup performance by Glasgow Coma Scale (GCS).

**Results:** Nineteen patients contributed 731 pupillometry recordings, of which 634 (87%) had concurrent invasive ICP measurements; 112 recordings (18%) showed elevated ICP. PuRe Score exhibited a significant inverse correlation with ICP (Spearman *ρ* =− 0.17, p<0.001). At the screening threshold (PuRe ≤1.3), sensitivity was 85.7% (95% CI: 78.0–91.0%) and specificity 61.3% (95% CI: 57.1–65.4%), yielding a negative predictive value of 95.2% (95% CI: 92.4–97.0%) with AUC of 0.72 (95% CI: 0.68–0.77). Mean ICP differed significantly across PuRe groups: unreactive pupils (PuRe=0) showed 18.3 *±*1.3 mmHg versus 8.2 ± 0.5 mmHg in brisk reactivity (PuRe 3; ≥ ANOVA F=24.27, p<0.001). The PuRe Score maintained stable discrimination across ambient lighting conditions (ANOVA p=0.91), whereas traditional constriction metrics showed significant lighting dependence (CAMP p=0.04, DELTA p=0.03).

**Conclusions:** In this pilot cohort, smartphone-based, lighting-invariant pupillometry demonstrated high negative predictive value for ruling out intracranial hypertension. A PuRe Score above the screening threshold may provide bedside reassurance that dangerous ICP elevation is unlikely, suggesting potential for a two-tier neuromonitoring strategy in which high-frequency non-invasive screening identifies patients who require targeted invasive monitoring. Larger validation studies are needed to confirm these findings.

## Introduction

### The Clinical Burden of Intracranial Hypertension

Intracranial hypertension is a time-sensitive driver of secondary brain injury across acute neurological emergencies, including traumatic brain injury, subarachnoid hemorrhage, intracerebral hemorrhage, ischemic stroke with edema, and hydrocephalus. When intracranial pressure (ICP) rises above compensatory thresholds, cerebral perfusion falls, tissue hypoxia ensues, and unchecked elevations lead to herniation syndromes and death. The relationship between sustained intracranial hypertension and poor outcome is well established: undetected or untreated ICP elevation compounds primary injury, making timely recognition central to critical care ^1,2^.

Invasive ICP monitoring—via intraparenchymal probes or ventricular catheters—remains the reference standard for quantifying intracranial dynamics and guiding targeted therapy. However, invasive monitoring requires neurosurgical expertise, specialized equipment, and exposes patients to procedural risks including hemorrhage, infection, and device malfunction ^3,4^. Consequently, uptake varies substantially across centers and injury types: in the international SYNAPSE-ICU cohort, ICP monitoring practice differed markedly between institutions, underscoring the gap between “reference standard” capability and real-world deployment ^2^. Many units reserve catheter placement for the sickest patients or those who have already deteriorated—precisely when earlier detection could have been most valuable.

### Non-Invasive Screening: A Pragmatic Alternative

A range of non-invasive strategies have been evaluated to identify elevated ICP, including neuroimaging surrogates, optic nerve sheath diameter (ONSD) ultrasonography, and transcranial Doppler (TCD) indices. Systematic reviews and meta-analyses indicate that single non-invasive modalities frequently lack the accuracy to replace invasive monitoring across heterogeneous populations ^5,6,7,8^. Even so, noninvasive methods can be clinically useful when positioned as high-sensitivity screening tools that rule out dangerous ICP elevation and guide escalation to invasive monitoring or imaging. This “screen-then-confirm” paradigm has been tested for TCD in brain-injured populations: the IMPRESSIT-2 multicenter study demonstrated that ultrasound-based screening could effectively exclude intracranial hypertension, reflecting a practical direction for scalable neuromonitoring pathways ^9^.

### The Penlight Examination: Limitations of Subjective Assessment

Pupillary size and reactivity are deeply embedded in neurological examination and ICP decision-making. Historically, these assessments have been performed manually with a penlight— an approach that is subjective, vulnerable to inter-observer variability, and poorly suited to detecting subtle changes over time ^10,11^. Multiple studies have demonstrated clinically meaningful disagreement in manual pupillary assessments, particularly for subtle reactivity changes that may herald evolving intracranial pathology. The penlight cannot provide quantitative metrics, cannot track trends reliably, and offers no objective documentation for longitudinal comparison.

### Infrared Pupillometry: A Partial Solution

Quantitative pupillometry was introduced to standardize the pupillary examination, providing objective, repeatable measurements of pupil diameter, constriction amplitude, velocities, latency, and recovery time ^12,13,14^. Composite indices aggregate these parameters into scalar scores that can be tracked over time. Across traumatic brain injury, intracerebral hemorrhage, and subarachnoid hemorrhage, depressed pupillary reactivity measured by quantitative pupillometry has been associated with intracranial hypertension, impending herniation, and poor outcome ^15,16,17,18,19,20^.

Across studies, diagnostic utility is often asymmetric: pupillometry is generally stronger for ruling out dangerous ICP elevation (high negative predictive value when indices are “normal”) than for ruling in elevated ICP (more limited positive predictive value). In an intracerebral hemorrhage cohort, Giede-Jeppe et al. reported high NPV for excluding intracranial hypertension, with limited PPV ^17^. This asymmetry has important clinical implications: a metric with high NPV can provide bedside reassurance that intracranial hypertension is unlikely, supporting a screening role even when the ability to confirm elevated ICP is modest.

### Limitations of Conventional Infrared Devices

Despite their promise, conventional infrared pupillometers have practical limitations. They rely on proprietary hardware with significant acquisition costs, are typically tethered to controlled lighting conditions, and remain susceptible to ambient light fluctuations ^21,22,23^. Recent studies in critically ill populations demonstrate that changes in room illumination can significantly shift composite pupillometry indices, potentially reclassifying patients across clinical thresholds ^21,22^. This light dependence complicates longitudinal interpretation, constrains deployment outside well-controlled ICU environments, and limits use in bright operating theatres, emergency departments, and pre-hospital settings.

### Computational Pupillometry: Leveraging Modern Hardware and AI

To address these gaps, recent work has introduced computational, lighting-invariant pupillometry. Modern smartphones equipped with neural processing units (such as the Apple Neural Engine) can execute advanced deep learning algorithms in real time, leveraging high-resolution cameras, precise LED flash control, and comprehensive sensor arrays including ambient light sensors and accelerometers. A machine-learning approach for ambient-light-corrected parameters and the Pupil Reactivity (PuRe) Score has demonstrated that a smartphone-based system can generate a scalar reactivity metric stable across four orders of magnitude in ambient illumination while discriminating reactive from non-reactive pupils with high accuracy ^24,25^.

Smartphone-based pupillometry has shown feasibility and diagnostic utility in TBI-related contexts, including clinician interpretation of smartphone PLR curves and classification tasks comparing smartphone parameters against commercial pupillometry outputs ^26,27^. However, not all smartphone implementations achieve robust agreement with established methods; iterative design refinement is required to ensure clinical-grade performance ^28^. The potential for deployment in resource-limited settings adds further motivation for developing validated, accessible platforms ^29,30^.

The conceptual framework for this study is illustrated in Figure 1. Invasive ventricular catheters (Figure 1A) remain the reference standard when neurosurgical resources are available, but a mobile, AI-enabled alternative (Figure 1B) could extend quantitative neuromonitoring to settings where invasiveness, staffing, or infection risk preclude catheter placement. The smartphone-based pupillometry enables bedside assessment without specialized equipment.

**Figure 1.**
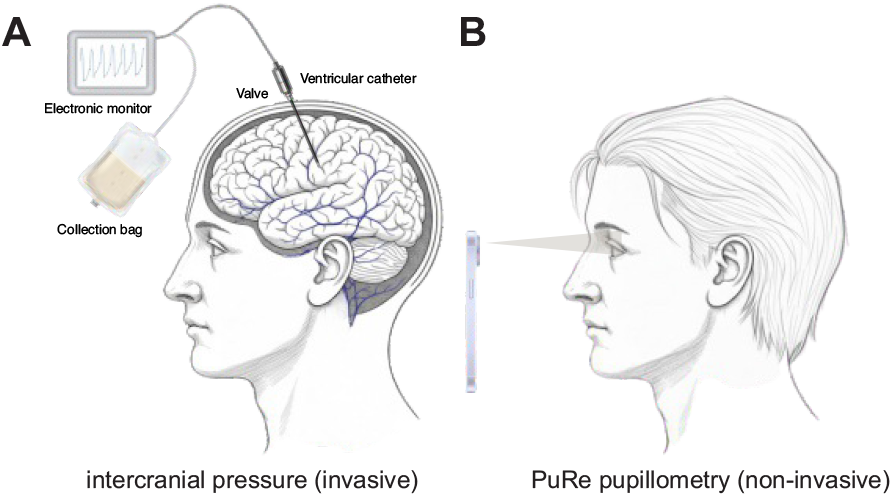
Conceptual framework of intracranial pressure monitoring strategies. (A) Invasive intracranial pressure monitoring using intraparenchymal probes as the reference standard. (B) Computational, lighting-invariant pupillometry as a non-invasive screening modality.

### Study Rationale and Objectives

Whether a mobile, lighting-corrected reactivity score can track invasive ICP in real-world intensive care, and whether it preserves the high negative predictive value that makes traditional pupillometry attractive as a screening tool for intracranial hypertension, has not been established. By focusing on a mobile, lighting-corrected metric validated against the invasive ICP reference standard, this study aims to inform a potential shift in neuromonitoring strategy: from sparse, invasive measurements in a minority of patients to highfrequency, non-invasive screening across the entire neuro-ICU population.

The primary objective was to determine the diagnostic performance of the lighting-invariant PuRe Score for identifying intracranial hypertension (ICP ≥ 20 mmHg), with emphasis on negative predictive value.

Secondary objectives were to: (i) quantify the correlation between PuRe Score and contemporaneous invasive ICP; (ii) characterize ICP burden across PuRe-defined reactivity strata; (iii) test whether the PuRe Score retains its performance across naturally variable ambient-light levels; and (iv) evaluate clinical associations with survival and neurological status.

All non-standard abbreviations are defined at first use.

## Methods

### Study Design and Oversight

This prospective, single-center pilot study was conducted in the neurological intensive care unit (neuro-ICU) of Gdańsk BJ Hospital, Department of Anesthesiology and Intensive Care (Gdańsk, Poland). The protocol adhered to the Declaration of Helsinki and received approval from the Bioethics Committee for Scientific Research at Gdańsk Medical University (KB/240/2025; 9 May 2025). Consecutive patients were enrolled between April and November 2025, and bedside follow-up continued until ICU discharge or death. This pilot study was designed to characterize the performance of a lighting-invariant quantitative pupillometry system relative to concurrently measured intracranial pressure (ICP), and reporting follows STROBE recommendations.

### Setting and Participants

The neuro-ICU is a tertiary, mixed medical–surgical service that manages acute stroke, traumatic brain injury, neurosurgical emergencies, and medical causes of intracranial hypertension. Eligible participants were adult patients (≥ 18 years) admitted with acute intracranial pathology and enrolled within 24 h of admission to the Department of Anesthesiology and Intensive Care. Inclusion diagnoses were: (i) traumatic brain injury with Glasgow Coma Scale (GCS) ≤12; (ii) ischemic stroke with respiratory failure; (iii) intracerebral hemorrhage with National Institutes of Health Stroke Scale (NIHSS) ≥ 5; and (iv) subarachnoid hemorrhage with Hunt–Hess grade *>*1. Exclusion criteria were: (i) history of major ocular surgery; (ii) retinal disease; (iii) ocular prosthesis; and (iv) acute brainstem pathology with anatomical involvement affecting pupillary reactivity.

Quantitative pupillometry was performed serially from admission day zero. Invasive ICP monitoring was performed when clinically indicated, and ICP-related analyses were restricted to recordings with concurrent ICP values. Due to the acute nature of the neurological conditions studied, the majority of patients were unable to provide informed consent at the time of enrolment because of impaired consciousness or ongoing analgosedation. The study protocol, including enrolment without prior consent, was reviewed and approved by the local Bioethics Committee. Deferred consent procedures were applied whenever possible, and informed consent was subsequently obtained from the patient once decision-making capacity was regained or from a legally authorized representative. Nineteen patients meeting eligibility criteria were enrolled, contributing 731 pupillometry recordings across 365 bedside sessions.

### Data Acquisition and Devices

All pupillary examinations were performed with the PuRe Pupillometer (Solvemed Inc., Lewes, DE, USA) operating on iPhone (Apple, Cupertino, CA, USA). The device leverages the phone’s camera, high-intensity LED flash, and dedicated neural processing unit to execute computer-vision pipelines in real time. Each acquisition produces raw pupil-diameter traces (60 frames per second) for both eyes together with a comprehensive feature set: initial diameter (INIT), constriction amplitude (CAMP), percent constriction (DELTA), constriction and dilation velocities, latency, recovery time, final diameter, symmetry metrics, and scene metadata including ambient illumination.

The Pupil Reactivity (PuRe) Score is a lighting-invariant metric derived from machine learning models that correct for ambient light and camera exposure effects, as described previously ^24^. Scores range from 0 to 5: values of 0 indicate unreactive pupils, 0.1–2.9 indicate sluggish reactivity, and ≥3 indicate brisk reactivity.

### Intracranial Pressure Monitoring

Intracranial pressure (ICP) was monitored using an intraparenchymal microstrain-gauge sensor with skull bolt fixation (Codman Microsensor metal skull bolt kit, REF 626638; Integra LifeSciences, Mansfield, MA, USA). The device was inserted by a neurosurgeon under sterile conditions according to the manufacturer’s instructions. After zeroing the transducer to atmospheric pressure in sterile fluid outside the body, a twist-drill burr hole was created and the metal skull bolt was screwed into the skull. The dura was perforated with the dedicated obturator, and the Microsensor catheter was advanced through the bolt so that the tip lay within the cerebral parenchyma (typically frontal white matter), guided by the depth markings on the catheter. The compression cap of the bolt was then tightened to secure the catheter and provide a CSF-tight seal. The sensor cable was connected to the bedside monitoring system, which provided continuous real-time ICP values and waveforms. ICP was monitored throughout the period of neuromonitoring as clinically indicated, and values were documented in the electronic medical record at regular intervals and around relevant clinical events (e.g., changes in sedation, osmotherapy, or surgical interventions). The device is a single-use, factory-calibrated solid-state transducer; no in-vivo recalibration was performed after implantation, and catheters were removed when ICP monitoring was no longer required or earlier if infection or device malfunction was suspected.

### Clinical Variables and Outcomes

Clinical data were captured in a unified prospectively maintained table that linked each recording to the patient’s Glasgow Coma Scale (GCS) score, mean arterial pressure (MAP), cerebral perfusion pressure (CPP = MAP – ICP), sedation or vasoactive infusions, survival status, and admission diagnosis. Demographic fields included age, sex, and ICU length of stay. ICP elevation was defined a priori as ICP≥ 20 mmHg based on institutional treatment thresholds.

### Recording Protocol and Quality Control

Recordings were obtained with the phone positioned approximately 15 cm from the examined eye. Each acquisition (60 Hz) consisted of a 1 s baseline, 1 s controlled flash stimulus, and 3 s recovery period. Both eyes were interrogated sequentially. Ambient light was measured with the device’s embedded algorithms, capturing the natural range observed in the ICU (approximately 10–1,000 lux, corresponding to log_10_ values of 1.0–3.0).

### Statistical Analysis

Analyses were conducted in Python 3.13 using <monospace> numpy, pandas, scipy, </monospace> and <monospace> scikit-learn.</monospace> Continuous variables are summarized as median with interquartile range (IQR) or mean ± standard error of the mean (SEM). Patients were stratified by PuRe Score (0, 0.1–2.9, ≥ 3.0) for descriptive comparisons. Between-group differences were assessed using one-way ANOVA with effect sizes (eta-squared) and Kruskal– Wallis tests with post-hoc procedures. Pearson and Spearman correlations quantified relationships between pupillometry parameters and clinical variables.

Diagnostic utility for detecting ICP ≥ 20 mmHg was explored with receiver operating characteristic (ROC) curves. Performance metrics (area under the ROC curve [AUC], NPV, sensitivity, specificity, PPV) are reported with 95% Wilson confidence intervals. A simple PuRe threshold was evaluated as a screening tool for elevated ICP.

The completed STROBE checklist for observational studies is provided as Supplementary Material.

## Results

### Study Cohort

Between April and November 2025, we enrolled 19 consecutive patients in the neurological intensive care unit who met eligibility criteria. Baseline characteristics of the study cohort are summarised in Table 1. The cohort included 17 males (89%) and 2 females (11%), with a median age of 53 years (IQR: 48–62). Primary diagnoses were subarachnoid haemorrhage (n=10, 53%) and traumatic brain injury (n=8, 42%), with one patient having other causes (n=1, 5%).

Disease severity at admission was substantial: the median Glasgow Coma Scale (GCS) was 5 (IQR: 4–11), with 68% of patients having severe neurological impairment (GCS≤ 8). For patients with subarachnoid haemorrhage, the median Hunt-Hess score was 3.5 (IQR: 3–5) and median Fisher score was 4 (IQR: 4–4). For patients with traumatic brain injury, the median Marshall CT classification was 5.5 (IQR: 4.5–6). During follow-up, 11 patients (58%) did not survive and 8 patients (42%) survived.

A total of 731 pupillometry recordings were obtained across 365 bedside sessions, with a median of 41 recordings per patient (IQR: 36–45). Intracranial pressure measurements were available for 634 recordings (87%), with 112 (18%) showing elevated ICP (≥20 mmHg).

### Intracranial Pressure and Pupillary Reactivity

We analyzed the relationship between intracranial pressure and pupillary light reflex characteristics in recordings with concurrent ICP measurements. Pupillary reactivity demonstrated a clear dose-response relationship with ICP severity (Figure 2). The mean PuRe Score progressively decreased from 2.03 in recordings with normal ICP (<20 mmHg) to 0.70 in those with moderately elevated ICP (20–40 mmHg), and further to 0.31 in recordings with severely elevated ICP (*>*40 mmHg). Representative averaged pupillograms (Figure 2A–C) illustrate the progressive dampening of the pupillary light reflex with increasing ICP, with high ICP recordings showing minimal constriction amplitude and markedly dilated baseline pupil diameter. Quantitative comparisons of pupillometry parameters across ICP categories (Figure 2D–G) confirmed these observations, with PuRe Score (Figure 2G) showing the clearest separation between ICP groups.

**Figure 2.**
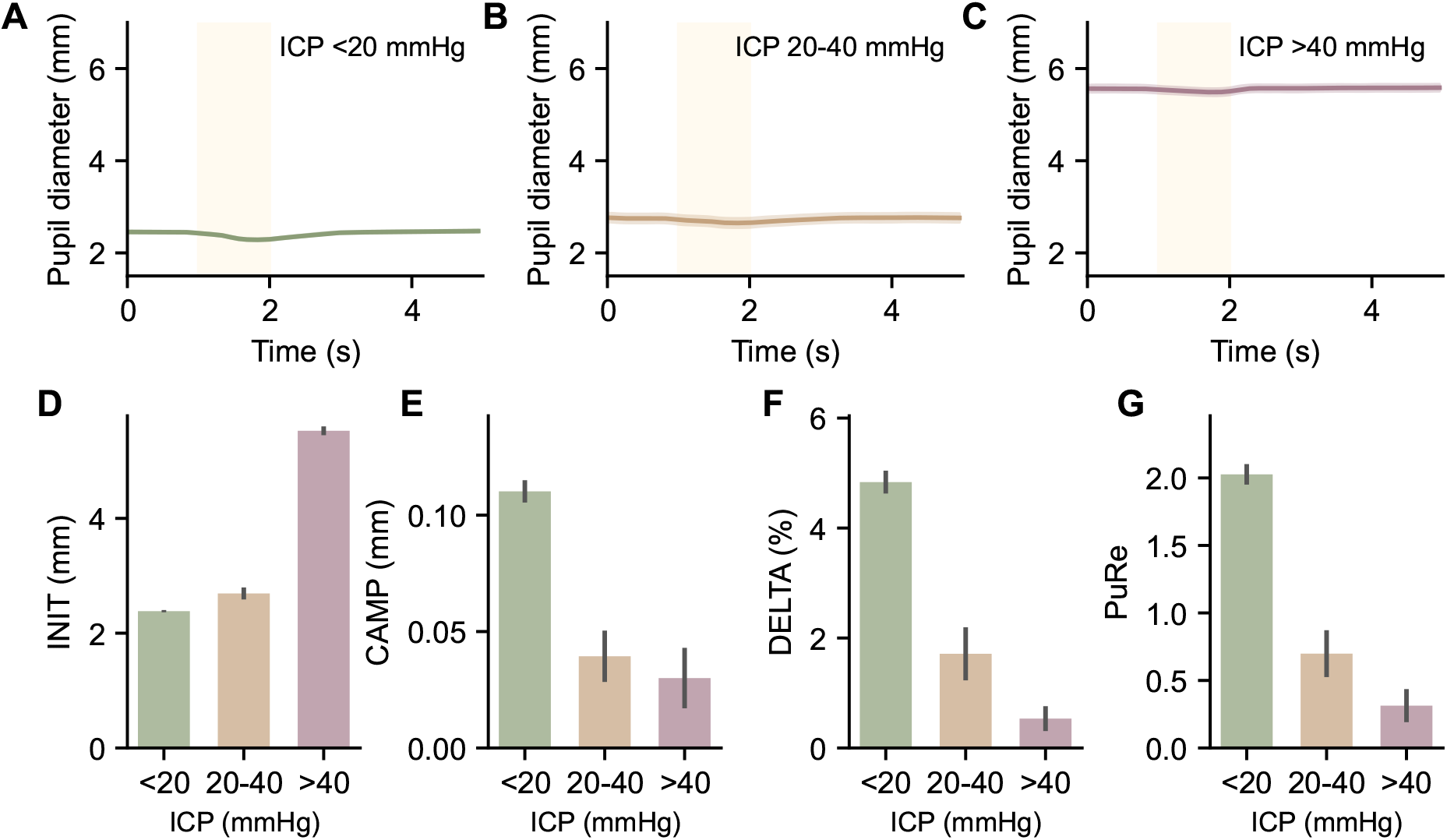
Relationship between pupillary reactivity and intracranial pressure. (A) Normal ICP (<20 mmHg), (B) moderately elevated ICP (20–40 mmHg), and (C) severely elevated ICP (*>*40 mmHg) averaged pupillograms. (D) Initial pupil diameter (INIT), (E) constriction amplitude (CAMP), (F) percent constriction (DELTA), and (G) Pupil Reactivity (PuRe) Score across ICP categories.

### Lighting Invariance of PuRe Score

A key advantage of the PuRe Score is its stability across varying ambient lighting conditions. We compared pupillometry parameters across three ambient light intensity categories: dark (<25 lux), medium (25–400 lux), and bright (*>*400 lux). As shown in Figure 3, traditional pupillometry parameters showed significant variation with ambient lighting: constriction amplitude (CAMP) varied significantly across lighting conditions (one-way ANOVA, F=3.19, p=0.04; Figure 3B), as did percent constriction (DELTA; F=3.37, p=0.03; Figure 3C). Initial pupil diameter (INIT) showed no significant variation (F=1.37, p=0.25; Figure 3A). Notably, the PuRe Score remained stable across all lighting conditions (F=0.10, p=0.91; Figure 3D), demonstrating its lighting-invariant design. This lighting invariance is essential for reliable pupillometry in variable clinical environments.

**Figure 3.**
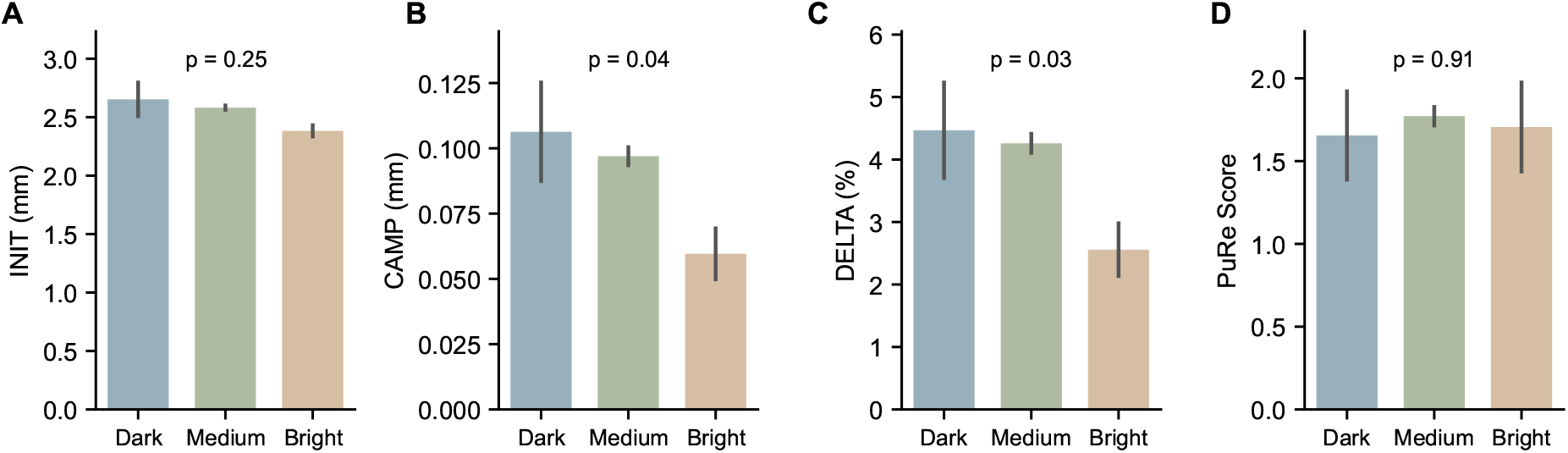
Effect of ambient lighting on pupillometry metrics. (A) Initial pupil diameter (INIT), (B) constriction amplitude (CAMP), (C) percent constriction (DELTA), and (D) Pupil Reactivity (PuRe) Score across three ambient light intensity categories (dark, medium, bright). Traditional pupillometry parameters demonstrate significant lighting dependence, whereas the PuRe Score remains stable across lighting conditions.

### Clinical Associations and Diagnostic Performance

The PuRe Score showed associations with clinical outcomes (Figure 4). Patients who did not survive had significantly lower PuRe Scores compared to survivors (1.53 ± 0.09 vs. 2.07 ± 0.09; p<0.001; Figure 4A). PuRe Scores were lower in recordings with severe neurological impairment (GCS ≤8) than in non-severe status (1.69 ± 0.07 vs. 2.04 ± 0.17), but this difference did not reach statistical significance in this cohort (p=0.06; Figure 4B), likely due to the inclusion criteria homogenizing the cohort.

**Figure 4.**
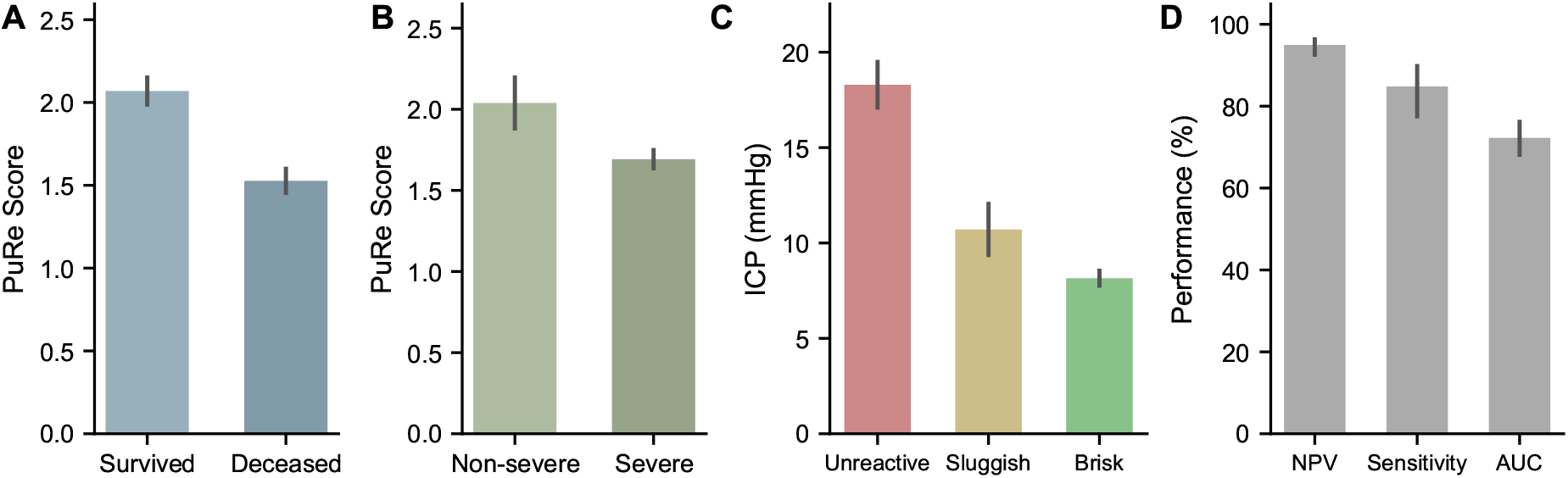
Clinical associations and diagnostic performance of the PuRe Score. (A) PuRe Score in survivors versus non-survivors. PuRe Score by Glasgow Coma Scale (GCS) category (severe GCS ≤8 vs. non-severe GCS*>*8). (C) Intracranial pressure (ICP) across PuRe reactivity strata (unreactive, sluggish, brisk). (D) Receiver operating characteristic (ROC) curve for detection of intracranial hypertension (ICP≥20 mmHg).

Mean ICP differed significantly across PuRe groups (Figure 4C). Recordings with unreactive pupils (PuRe=0) had elevated ICP (18.3 ± 1.3 mmHg), compared to those with sluggish (0<PuRe<3; 10.7 ± 1.4 mmHg) or brisk reactivity (PuRe *≥*3; 8.2± 0.5 mmHg; one-way ANOVA F=24.27, p<0.001, *η*^2^=0.071).

For predicting elevated ICP (≥ 20 mmHg), the PuRe Score at a screening threshold of PuRe *≤* 1.3 demonstrated high negative predictive value (NPV=95.2%, 95% CI: 92.4–97.0%) and sensitivity (85.7%, 95% CI: 78.0–91.0%), with an area under the receiver operating characteristic curve (AUC) of 0.72 (95% CI: 0.68–0.77) (Figure 4D). These results indicate that a reactive pupil (PuRe*>*1.3) effectively rules out elevated ICP, with only 16 of 336 recordings (4.8%) with PuRe*>*1.3 having elevated ICP.

### Recording-Level Correlations

Recording-level correlations confirmed that the PuRe Score tracked invasively measured ICP (Spearman *ρ* = − 0.17, p<0.001). This inverse relationship was consistent across diagnostic subgroups: PuRe was significantly lower in SAH patients compared to TBI patients (t=-2.11, p=0.04, Cohen’s d=-0.16), while ICP did not differ significantly between groups (p=0.15). Patient-level analyses showed that lower mean PuRe Score was associated with mortality (AUC=0.69 for predicting death), though this did not reach statistical significance in univariate comparison (p=0.23).

## Discussion

### Principal Findings

In this pilot cohort, a smartphone-based computational pupillometry system produced a lighting-invariant Pupil Reactivity (PuRe) Score that tracked invasive ICP in clinically meaningful ways. The PuRe Score showed a significant inverse correlation with ICP (Spearman *ρ* = − 0.17, p<0.001) and a clear dose–response across reactivity strata: recordings with absent reactivity (PuRe=0) coincided with the highest ICP values (18.3 ± 1.3 mmHg), whereas brisk reactivity (PuRe ≥ 3) aligned with near-normal pressures (8.2 ± 0.5 mmHg; ANOVA F=24.27, p<0.001, *η*^2^=0.071).

At the screening threshold (PuRe ≤1.3), the negative predictive value reached 95.2% (95% CI: 92.4–97.0%), suggesting that recordings above this threshold rarely coincided with intracranial hypertension (ICP ≥ 20 mmHg). This high NPV, combined with lighting-invariant performance and mobile deployment, suggests the PuRe Score may serve as a practical screening tool for ruling out dangerous ICP elevation.

### Relation to Prior Pupillometry–ICP Evidence

The observed pattern—moderate correlation with ICP but strong NPV for excluding intracranial hypertension—mirrors prior work using dedicated infrared pupillometers and composite indices. Multiple cohorts report that preserved quantitative reactivity is rarely present during sustained intracranial hypertension, producing strong “rule-out” performance even when the relationship between any single measurement and concurrent ICP is noisy ^15,16,17,18,20^. McNett et al. found significant correlations between hourly pupillometer readings and ICP values, supporting the physiological linkage between pupillary dynamics and intracranial pressure ^15^.

In intracerebral hemorrhage, Giede-Jeppe et al. specifically used automated pupillometry to identify the absence of ICP elevation, reporting high NPV ^17^. Our pilot findings are consistent with this prior work and preliminarily extend the principle to a smartphone-based, computationally corrected platform. The recent ORANGE program established the prognostic value of quantitative pupillometry trajectories in severe non-anoxic acute brain injury ^19^, and secondary analyses linked pupillometry indices with intracranial hypertension risk ^20^—supporting pupillometry as a meaningful physiologic signal even when correlation with instantaneous ICP is not tight.

It is also consistent with broader physiology that the pupil light reflex reflects an integrated brainstem and autonomic response rather than a direct manometer. Instantaneous ICP is influenced by interventions, patient positioning, ventilatory mechanics, and short-time-scale dynamics; pupillary metrics are additionally shaped by sedatives, analgesics, and baseline ocular characteristics. Therefore, modest point-by-point correlation (*ρ* = − 0.17) does not preclude strong screening utility, particularly when the goal is to detect (or confidently exclude) clinically meaningful ICP elevations at the bedside.

### Screening Threshold and Clinical Implications

At the PuRe ≤1.3 threshold, performance favors sensitivity (85.7%) over specificity (61.3%). In practical terms, this means that most clinically important ICP elevations fall into the low-PuRe range, while episodes of ICP ≥ 20 mmHg with PuRe*>*1.3 were uncommon (only 16 of 336 recordings, 4.8%). This asymmetry is clinically appropriate: the consequences of a missed hypertensive episode are severe, but the downside of false-positive alerts is relatively low—prompting additional evaluation rather than irreversible intervention.

### Two-Tier Neuromonitoring Strategy

These pilot results suggest a potential two-tier neuromonitoring model aligned with broader non-invasive neuromonitoring reviews ^3,5,6^. The first tier involves high-frequency, non-invasive screening using mobile, lighting-invariant pupillometry across the broader neuro-ICU population. Persistently normal or improving PuRe Scores (PuRe*>*1.3) may provide reassurance that dangerous intracranial hypertension is unlikely. The second tier involves targeted invasive monitoring for patients with depressed or deteriorating PuRe Scores, new asymmetries, or discordant clinical features. In these patients, invasive ICP confirms and quantifies the burden of intracranial hypertension and guides therapy.

This mirrors the “screen-then-confirm” paradigm tested for TCD-based ICP assessment in the IMPRESSIT-2 study ^9^. It also harmonizes with how pupillometry has been positioned in prior work: a bedside signal with strong negative predictive properties and value for trend detection rather than a standalone surrogate for absolute ICP.

Instantaneous ICP and pupillary reactivity reflect related but non-identical physiological processes, and modest point-wise correlation does not preclude strong screening performance for clinically meaningful ICP thresholds.

In practice, such a strategy, if validated in larger cohorts, could (i) broaden surveillance to patients who would not otherwise receive an ICP catheter, (ii) provide earlier warning to prompt clinician reassessment or imaging, and (iii) reduce unnecessary invasive monitoring in patients with persistently reassuring reactivity trajectories—though the latter requires careful prospective evaluation to ensure that “reassuring” screens do not delay intervention in atypical presentations.

### Lighting Invariance and Clinical Robustness

Ambient light is an underappreciated confounder in pupillometry. Conventional devices often require darkened rooms or shielded optics to reduce variance ^21,22,23^. Studies in healthy volunteers and critically ill populations demonstrate that lighting conditions can shift multiple pupillary parameters, and even when changes in composite indices may be “small on average,” they can be inconsistent and may reclassify patients across clinical thresholds ^21,22^.

By comparison, the PuRe Score showed no significant dependence on ambient lighting (ANOVA p=0.91), whereas traditional constriction metrics varied significantly (CAMP p=0.04, DELTA p=0.03). This lighting robustness is not a minor ergonomic point: it reduces operational burden (no need to darken rooms or shield optics), improves longitudinal comparability of repeated measurements, and expands potential use contexts—including bright operating theatres, emergency departments, and pre-hospital settings where traditional devices are constrained. If replicated across centers and hardware variants, this feature could shift pupillometry from “occasional device-based test” to “routine, high-frequency physiologic vital sign” in acute brain injury pathways.

### Smartphone Platform Considerations

Smartphone-based pupillometry is increasingly supported by clinical validation work in TBI-adjacent contexts. McGrath et al. demonstrated that clinicians could interpret smartphone PLR curves for TBI diagnosis ^26^, and Maxin et al. validated a smartphone pupillometry application against commercial devices for severe TBI classification ^27^. This pilot study contributes by providing initial validation against the invasive ICP reference standard (rather than against another pupillometer) and by preliminarily optimizing for a screening goal (maximizing sensitivity and NPV).

However, not all smartphone implementations achieve robust performance. Tan et al. reported an unsuccessful prototype and analyzed the causes of failure, emphasizing that iterative design refinement is essential for clinical-grade reliability ^28^. The potential for deployment in resource-limited settings adds urgency to developing validated, accessible platforms ^29,30^.

### Integration with Multimodal Monitoring

An important future direction is the integration of computational pupillometry with other non-invasive neuromonitoring signals. The IMPRESSIT-2 study demonstrated that TCD can screen for intracranial hypertension ^9^, and ONSD ultrasonography has been evaluated in multiple meta-analyses ^7,8^. Combining pupillometry with TCD, ONSD, or other physiologic indices could improve discrimination beyond what any single modality achieves, potentially enabling more confident triage decisions in settings where invasive monitoring is unavailable or undesirable.

### Clinical Associations

Beyond ICP prediction, the PuRe Score showed associations with clinical outcomes. Patients who did not survive had lower PuRe Scores than survivors (1.53 vs. 2.07, p<0.001), while PuRe Scores were lower in recordings with severe neurological impairment (GCS ≤8) than in non-severe status (1.69 vs. 2.04) but did not reach statistical significance (p=0.06). While the correlation between PuRe and continuous ICP was modest (*ρ* =− 0.17), the strong association with categorical ICP elevation (normal vs. elevated) and clinical outcomes suggests potential utility as a neuromonitoring adjunct. These pilot findings align with the ORANGE program, which established prognostic value of pupillometry indices in acute brain injury ^19^.

### Limitations

This pilot study has important limitations that should guide interpretation and future work.

First, this is a single-center pilot study with a modest number of patients (n=19). The small sample size and single-center design limit generalizability and require validation in larger, multicenter cohorts. The cohort studied consisted mainly of men, which was a coincidence – during the study period, it was precisely these patients who were admitted consecutively, so repeated measures and local practice patterns may influence effect sizes and limit generalizability. The cohort consisted of neuro-ICU patients already selected for invasive monitoring, which can enrich for severe physiology and may not represent emergency department, ward, or pre-hospital populations.

Second, the analysis is largely at the recording level (731 recordings, 634 with paired ICP), which inflates the apparent sample size; patient-level clustering was addressed through stratified analyses but not formal mixed-effects modelling. Future validation should consider hierarchical models and patient-level endpoints to quantify robustness under within-subject dependence.

Third, NPV estimates depend on event prevalence: the 18% prevalence of ICP ≥ 20 mmHg in this cohort may differ from other populations. If implemented in settings with lower prevalence (e.g., broader emergency populations), NPV would typically increase while PPV decreases; conversely, in highly selected cohorts with higher prevalence, NPV could decline. For clinical translation, future studies should report likelihood ratios, calibration across prevalence strata, and patient-level decision performance.

Fourth, we did not systematically characterize ophthalmic comorbidities or sedative dosing. While these factors do not negate screening utility, they can affect thresholds and false-positive rates. Stratified analyses by sedation class and ocular pathology would strengthen clinical interpretability.

Finally, this work focused on short-term physiological endpoints; longer-term functional outcomes were not incorporated.

### Future Directions

Future research should address several key questions. First, can PuRe-based thresholds, when embedded into predefined neuromonitoring algorithms, safely guide decisions about ICP monitoring in larger, multicenter cohorts? Second, can the integration of computational pupillometry with other non-invasive signals (TCD, ONSD) improve discrimination? Third, how does mobile pupillometry perform in pre-hospital, emergency, and intraoperative environments? Fourth, what is the patient-level false-negative rate—that is, how often does a “reassuring” PuRe trajectory precede a clinically significant ICP crisis?

## Conclusions

In this pilot intensive care study, a smartphone-based, lighting-invariant pupillometry system demonstrated a negative predictive value of 95.2% for ruling out intracranial hypertension at the PuRe Score screening threshold (≤1.3). The PuRe Score maintained stable performance across variable ambient lighting conditions and showed strong associations with ICP severity strata and clinical outcomes.

These pilot findings suggest computational, mobile pupillometry may serve as a high-NPV screening tool for intracranial hypertension, consistent with the “rule-out” utility observed with conventional infrared devices but without the constraints of proprietary hardware and controlled lighting. Rather than competing with invasive ICP monitoring, such systems, if validated in larger studies, could enable a two-tier neuromonitoring strategy: high-frequency, lighting-invariant reactivity measurements guide which patients require the risks and resources of invasive monitoring and which can be safely managed without it. Larger, multicenter validation studies are needed to confirm these preliminary findings and establish generalizability across diverse patient populations and clinical settings.

## Data Availability

The data that support the findings of this study are available from the corresponding author upon reasonable request. The data are not publicly available due to privacy concerns for the research participants.

## Additional Information

### Author Contributions

K.C. and M.W. conceived the study. K.C., M.Wu., A.F., R.M.R., G.G., J.K., P.K., and K.S. contributed to patient enrollment and data acquisition. J.C., M.S., H.C., M.D., and R.C. contributed to software development, data processing, and quality control. M.W. and R.C. performed statistical analyses. K.C. and M.W. drafted the manuscript. All authors critically revised the manuscript for important intellectual content and approved the final version.

### Authorship Confirmation

All authors meet the International Committee of Medical Journal Editors (ICMJE) criteria for authorship and approve the final manuscript.

### Ethical Approval

The study was approved by the Bioethics Committee for Scientific Research at the Medical University of Gdańsk (approval number KB/240/2025). The study was conducted in accordance with the Declaration of Helsinki and its later amendments.

### Informed Consent

Due to impaired consciousness in the majority of patients at enrollment, informed consent was deferred. Consent was subsequently obtained from the patient when capacity returned or from a legally authorized representative, in accordance with institutional and ethical committee approval.

### Statement of Human and Animal Rights

This study involved human participants only. All procedures were performed in accordance with institutional and national ethical standards. No animal studies were performed.

### Reporting Guidelines

This study was conducted and reported in accordance with the STROBE (Strengthening the Reporting of Observational Studies in Epidemiology) guidelines. The completed STROBE checklist is provided as a supplementary file.

